# Quantitative assessment of motor neglect

**DOI:** 10.1101/2020.07.01.20144170

**Authors:** Monica N. Toba, Chiara Pagliari, Marco Rabuffetti, Norbert Nighoghossian, Gilles Rode, François Cotton, Lucia Spinazzola, Francesca Baglio, Raffaella Migliaccio, Paolo Bartolomeo

## Abstract

**Objectives:** We used differential actigraphy as a novel, objective method to quantify motor neglect (a clinical condition whereby patients mimic hemiplegia even in the absence of sensorimotor deficits), whose diagnosis is at present highly subjective, based on the clinical observation of patients’ spontaneous motor behavior.

**Methods:** Patients wear wristwatch-like accelerometers, which record spontaneous motor activity of their upper limbs during 24 hours. Asymmetries of motor behavior are then automatically computed offline. On the basis of normal participants’ performance, we calculated cut-off scores of left/right motor asymmetry.

**Results:** Differential actigraphy showed contralesional motor neglect in nine of 35 patients with unilateral strokes, consistent with clinical assessment. An additional patient with clinical signs of motor neglect obtained a borderline asymmetry score. Lesion location in a subgroup of 25 patients was highly variable, suggesting that motor neglect is a heterogenous condition.

**Conclusions:** Differential actigraphy provides an ecological measure of spontaneous motor behavior, and can assess upper limb motricity in an objective and quantitative manner. It thus offers a convenient, cost-effective, and relatively automatized procedure for following-up motor behavior in neurological patients, and to assess the effects of rehabilitation.

## INTRODUCTION

Patients with unilateral brain damage may avoid moving the limbs contralateral to their lesion, even in the absence of sensorimotor deficits [1]. However, these patients typically show normal strength and dexterity when asked to move their limbs. Laplane and Degos [2] dubbed this condition Motor Neglect (MN). They described 20 stroke patients with “pure” MN (12 patients with right hemisphere lesions, eight with left hemisphere lesions), without substantial sensorimotor deficits or signs of visual neglect. Subsequent studies reported MN in 12-33% of acute stroke patients and ∼8% of chronic stroke patients [3,4]. Typically, these patients tend to use the ipsilesional limb even when the use of the contralesional limb would be more appropriate and convenient. No or little involvement of the contralesional limb occurs in gesture during speaking and in bimanual tasks (e.g., clapping, opening a bottle, buttoning or unbuttoning a dress). During walking, the contralesional limb may lag behind the ipsilesional limb, or it may lack normal swinging. Also, the characteristics of contralesional limb movements can be anomalous: movements can be delayed (hypokinesia), slowed (bradykinesia), and of reduced amplitude (hypometria). MN can co-occur with personal neglect (inattention for the contralesional side of the body), or with visual neglect (inattention for the contralesional side of space) [5]. However, the patterns of association are unclear, also because personal neglect has rarely been assessed in MN patients [3]. In principle, impaired conscious processing of the contralesional half of the body could impact the representation for perception (resulting in personal neglect), the representation for action (resulting in MN), or both [6,7]. Also sensorimotor deficits can accompany MN: patients with mild hemiparesis may display less spontaneous movement than predicted by their elementary motor deficit [8].

Anatomically, MN can occur after lesions in either hemisphere. Intra-hemispheric sites of lesion include the medial frontal premotor and motor areas [2,9,10], medial parietal regions [2,10,11], putamen, internal capsule and the thalamus [2,9,10,12–14]. Lesion locations in the white matter include the corpus callosum, fronto-parietal connections [2,10], and the cingulum [15,16].

MN can be severely disabling, because in severe cases it can mimic hemiplegia. Assessing MN has thus important clinical implications for patient management and rehabilitation. However, MN diagnosis is at present exclusively clinical, based on the observation of patients’ spontaneous motor behavior. The present study had two aims: (1) introduce an objective and quantitative assessment method for MN, based on differential actigraphy, which provides continuous assessment of spontaneous movements over 24h [17]; (2) explore the lesional correlates of MN.

## MATERIALS AND METHODS

### Participants

We originally recruited 50 patients with a first unilateral stroke. Inclusion criteria were: preserved capacity to understand the test requirements, no severe general mental deterioration, absence of psychiatric disorders or prior history of neurological disease, preserved motor and sensory capacities in the ipsilesional upper limb, absence of elementary motor deficits other than hemiplegia or hemiparesis. Patients were recruited in three clinical centers: IRCCS Fondazione Don Carlo Gnocchi Milan, Neuropsychology Unit of the A. Bellini Hospital Somma Lombardo, and the Stroke Unit of the Pierre Wertheimer Neurological Hospital Lyon. Thirteen patients were subsequently excluded (presence of non-vascular lesions, bilateral lesions or lesions restricted to the cerebellum). Two additional patients (P10 and P25) were subsequently excluded from the analysis due to technical problems in actigraphy data acquisition. Therefore, 35 patients constituted the final sample: 21 men (mean age, 63.8 years; SD, 13; range 38-86 years), and 14 women (mean age, 52.2 years; SD, 14.9; range 23-74 years). Seven of these patients had left hemisphere lesions, 28 had right hemisphere lesions; 34 patients were right-handed at Edinburgh Handedness Inventory [18], one patient (P30) was left-handed. The mean time of testing since brain damage onset was 99 days (range 2-1859 days). Neuroimaging data were only available for 25 patients with right hemisphere lesions (mean time of MRI acquisitions since symptom onset, 167 days; range, 66-458 days). Table 1 reports patients’ demographical and clinical details.

**Table 1.** Demographical and clinical characteristics of patients. M, male; F, female; R, right; L, left; I, ischemic; H, haemorrhagic; **, severe motor asymmetry; *, moderate motor asymmetry. For the handedness score [18], positive values indicate right handedness, negative values indicate left handedness.

| Patient | Sex/Age/<br>Education<br>(years) | %<br>Handedness<br>(Edinburgh<br>inventory) | Aetiology | Side<br>of<br>lesion | Delay<br>since<br>onset<br>(days) | Neurological deficits |  | Motricity Index<br>Score<br>(normal=100)<br>L/ R | Upper Limb<br>Modified<br>Motricity Index<br>Score<br>(normal=0) |
| --- | --- | --- | --- | --- | --- | --- | --- | --- | --- |
|  |  |  |  |  |  | Visual Field | Somatosensory |  |  |
| P1 | M/86/5 | 80 | I | L | 7 | Normal | Normal | 100/100 | 0 |
| P2 | M/66/23 | 100 | I | R | 10 | Left<br>hemianopia | Left hemianesthesia | 10/100** | 90** |
| P3 | F/73/5 | 100 | I | R | 23 | Left<br>extinction | Normal | 10/100** | 90** |
| P4 | F/50/15 | 100 | I | R | 37 | Normal | Normal | 100/100 | 0 |
| P5 | F/23/12 | 100 | I | R | 6 | Normal | Normal | 84/100* | 16* |
| P6 | M/38/10 | 100 | I | R | 2 | Normal | Left hemianesthesia | 100/100 | 0 |
| P7 | F/34/12 | 100 | I | L | 6 | Normal | Normal | 100/100 | 0 |
| P8 | F/68/5 | 100 | I | L | 3 | Normal | Normal | 100/100 | 0 |
| P9 | M/38/12 | 100 | I | R | 11 | Normal | Left hemianesthesia | 10/100** | 90** |
| P11 | M/58/12 | 90 | I | R | 3 | Normal | Left hemianesthesia | 100/100 | 0 |
| P12 | M/65/12 | 100 | I | R | 6 | Normal | Normal | 100/100 | 0 |
| P13 | M/66/15 | 100 | I | R | 8 | Normal | Normal | 100/100 | 0 |
| P14 | M/50/12 | 100 | H | R | 2 | Left<br>hemianopia | Left hemianesthesia | 19/100** | 81** |
| P15 | M/63/9 | 100 | I | R | 2 | Normal | Normal | 100/100 | 0 |
| P16 | M/75/9 | 100 | I | R | 4 | Left<br>hemianopia | Left hemianesthesia | 100/100 | 0 |
| P17 | F/54/15 | 100 | I | R | 7 | Normal | Left hemianesthesia | 100/100 | 0 |
| P18 | M/74/12 | 100 | I | R | 2 | Normal | Normal | 100/100 | 0 |
| P19 | M/56/9 | 100 | I | R | 3 | Normal | Normal | 77/100* | 23* |
| P20 | F/59/12 | 90 | I | R | 6 | Left<br>extinction | Left hemianesthesia | 10/100** | 90** |
| P21 | F/41/12 | 100 | I | R | 5 | Normal | Normal | 15/100** | 85** |
| P22 | F/57/9 | 100 | H | L | 7 | Normal | Normal | 100/10** | -90** |
| P23 | M/86/5 | 70 | I | R | 4 | Normal | Normal | 40/100** | 60** |
| P24 | M/61/12 | 100 | I | R | 5 | Normal | Normal | 100/100 | 0 |
| P26 | F/42/12 | 90 | I | R | 8 | Left<br>hemianopia | Normal | 100/100 | 0 |
| P27 | F/62/12 | 80 | H | R | 3 | Normal | Normal | 100/100 | 0 |
| P28 | M/79/4 | 100 | I | L | 723 | Right<br>extinction | Right<br>hemianesthesia | 100/10** | -90** |
| P29 | M/72/8 | 100 | I | L | 1859 | Normal | Right<br>hemianesthesia | 100/10** | -90** |
| P30 | M/50/13 | -70 | I | L | 31 | Normal | Normal | 100/100 | 0 |
| P31 | F/52/13 | 100 | I | R | 338 | Normal | Normal | 73/100* | 27* |
| P32 | M/60/23 | 100 | H | R | 103 | Left<br>extinction | Left hemianesthesia | 100/100 | 0 |
| P33 | F/63/8 | 100 | I | R | 109 | Normal | Left hemianesthesia | 75/100* | 25* |
| P34 | F/74/13 | 90 | I | R | 41 | Left extinction | Left hemianesthesia | 10/100** | 90** |
| P35 | M/71/10 | 100 | I | R | 67 | Normal | Normal | 100/100 | 0 |
| P36 | F/41/16 | 100 | I | R | 111 | Normal | Normal | 34/100** | 66** |
| P37 | M/62/16 | 100 | H | R | 65 | Normal | Left hemianesthesia | 100/100 | 0 |

### Standard Protocol Approvals, Registrations, and Patient Consents

All patients gave written consent according to the Declaration of Helsinki. The study was previously approved by the Ethics Committee of the Don Gnocchi Foundation (approved on 9/04/2014).

### Neuropsychological assessment

Patients underwent the GEREN battery [19] for the assessment of visual neglect. The battery includes bells test, landscape drawing, line bisection, writing, identification of overlapping figures and clock drawing. Additionally, we administered tests of letter cancellation [20] and line cancellation [21]. Personal neglect was assessed by means of the Fluff test [22], the Comb & Razor test [23,24], and the Bisiach test [25]. We used the Catherine Bergego Scale [26] to evaluate anosognosia. We also assessed patients’ preferential gaze orientation [27] (Table 2).

**Table 2.** Neuropsychological results. *, pathological scores compared to normative data (for gaze orientation, see [27]; for anosognosia, see [26]; for Albert’s test, see [21]; for bells cancellation, writing, line bisection, landscape drawing, clock drawing and overlapping figures, see [19]; for the Fluff test, see [22]; for the Bisiach test, see [25]; for the Comb and Razor test, see [23-24]. NA, not available; NE, not evaluable.

| Patient | Gaze Orientation<br>(0=no deviation) | Anosognosia | Albert's Test<br>(left/right hits, max= 30/30) | Letter cancellation<br>(left/right hits, max= 30/30) | Bells cancellation<br>(left/right hits, max= 15/15) | Writing<br>(cm from left margin) | 200mm line bisection<br>(mm of rightward deviation) | Landscape drawing score<br>(elements omitted, max= 6) | Clock drawing<br>(contralesional omission, max= 2) | Overlapping figures<br>(left/right hits= 5/5) | Fluff Test<br>(targets omitted, max=9) | Bisiach Test<br>(closed eyes) | Comb and Razor Test |
| --- | --- | --- | --- | --- | --- | --- | --- | --- | --- | --- | --- | --- | --- |
| P1 | 0 | - | 30/30 | 30/30 | 15/15 | 0.6 | -0.5 | 0 | 0 | 5/5 | 0 | 0 | 1.8 |
| P2 | 1 | - | 30/30 | 13/30* | 4/13* | 11.3* | 3 | 0 | 1* | 5/5 | 5* | 0 | 7 |
| P3 | 3 | + | 0/23* | 6/29* | 0/14* | 9* | 5.5 | 0 | 2* | 5/5 | 10* | 0 | 36* |
| P4 | 0 | - | 30/30 | 30/30 | 14/15 | 2 | 0.5 | 0 | 0 | 5/5 | 0 | 0 | 9 |
| P5 | 0 | - | 30/30 | 29/29* | 13/12 | 11* | 3 | 0 | 0 | 5/5 | 0 | 0 | 4 |
| P6 | 0 | - | 30/30 | 30/30 | 15/15 | 9* | 1.5 | 0 | 0 | 5/5 | 1 | 1* | 5 |
| P7 | 0 | - | 30/30 | 30/30 | 13/14 | 3 | -3.5 | 1* | 0 | 5/5 | 0 | 0 | 2 |
| P8 | 0 | - | 30/30 | 30/30 | 13/15 | 4.2 | -1 | 0 | 0 | 5/5 | 0 | 0 | 8 |
| P9 | 0 | + | 30/30 | 29/28* | 14/14 | 3.5 | 8* | 0 | 0 | 5/5 | 4* | 0 | 27* |
| P11 | 0 | + | 30/30 | 30/30 | 13/15 | 6 | 1.5 | 0 | 0 | 5/5 | 0 | 0 | 1.9 |
| P12 | 0 | - | 29/30 | 29/20* | 13/15 | 9.2* | 7.5* | 0 | 0 | 5/5 | 1 | 0 | 1 |
| P13 | 0 | - | 30/30 | 30/29 | 14/14 | 4 | 0.5 | 0 | 0 | 5/5 | 4* | 0 | 1.5 |
| P14 | 3 | - | 28/30 | 0/27* | 0/12* | 10.5* | 13* | 1* | 0 | 4/4* | 2* | 1* | 28* |
| P15 | 0 | - | 30/30 | 30/30 | 15/15 | 2.5 | 6.5* | 0 | 0 | 5/5 | 0 | 0 | 1.3 |
| P16 | 0 | - | 30/30 | 26/23* | 14/15 | 5 | 4.5 | 0 | 0 | 5/5 | 0 | 0 | 1.6 |
| P17 | 0 | - | 30/30 | 30/29 | 15/15 | 3.5 | 9* | 0 | 0 | 5/5 | 1 | 0 | 3 |
| P18 | 0 | - | 30/30 | 30/30 | 14/14 | 6.5 | 0.5 | 0 | 0 | 5/5 | 0 | 0 | 7 |
| P19 | 1 | - | 30/30 | 30/30 | 12/15* | 3 | 14* | 1* | 0 | 5/5 | 0 | 0 | 9 |
| P20 | 3 | + | 29/30 | 18/30* | 11/13 | 7.5 | 5 | 0 | 0 | 4/4* | 8* | 1* | 34* |
| P21 | 1 | - | 30/30 | 29/30* | 15/14 | 7.2 | 6.5* | 0 | 1* | 5/5 | 1 | 1* | 1.9 |
| P22 | 0 | - | 30/30 | 30/30 | 13/12 | 3.6 | -4.5 | 0 | 0 | 5/5 | 0 | 0 | 2.4 |
| P23 | 0 | - | 30/30 | 29/30 | 13/15 | 9.5* | 2.5 | 0 | 0 | 5/5 | 0 | 1* | 4 |
| P24 | 1 | - | 30/29 | 30/30 | 15/15 | 2.5 | 4.5 | 0 | 0 | 5/5 | 0 | 0 | 2.9 |
| P26 | 1 | - | 30/30 | 30/30 | 15/15 | 1.2 | 8.5* | 0 | 0 | 5/5 | 0 | 0 | 3 |
| P27 | 0 | - | 30/30 | 30/30 | 13/13 | 2 | 1.5 | 0 | 0 | 5/5 | 0 | 0 | 6 |
| P28 | 0 | - | NA | NA | NA | 2.5 | NA | NA | NA | NA | 0 | 0 | NA |
| P29 | 0 | - | 29/29 | 27/27* | 13/12 | 2.1 | 0.1 | 0 | 0 | 5/5 | 0 | 0 | 4.4 |
| P30 | 0 | - | 30/30 | 30/29 | 15/15 | NA | -0.2 | 0 | NA | 5/5 | 0 | 0 | 1.5 |
| P31 | 0 | - | 26/29* | 26/27* | 13/12 | 2.5 | -0.8 | 1* | 0 | 5/5 | 0 | 0 | 3 |
| P32 | 2 | - | 27/30* | 30/30 | 15/14 | 4.7 | -0.1 | 0 | 0 | 5/5 | 1 | 0 | 10 |
| P33 | 0 | - | 16/17 | 24/20* | 14/12* | 9.5* | 1.8 | 1* | 2* | 5/5 | 3* | 0 | 27* |
| P34 | 0 | - | 27/30* | 18/17* | 8/9* | 6.7 | 4.7 | 2* | 0 | 4/5* | 2* | 0 | 3.2 |
| P35 | 0 | - | 30/30 | 30/30 | 15/15 | 2.5 | 0.1 | 0 | 0 | 5/5 | 0 | 0 | 2.3 |
| P36 | 0 | - | 30/30 | 28/28* | 15/11* | 9.5* | -0.5 | 0 | 0 | 5/5 | 0 | 0 | -23* |
| P37 | 1 | - | 30/30 | 23/30* | 12/15* | 8.3* | 5.5 | 3* | 0 | 5/5 | 0 | 0 | 8 |

### Clinical motor assessment

The Motricity Index [28] was used to clinically assess pinch grip, elbow flexion and shoulder abduction in the upper limb and ankle dorsiflexion, knee extension and hip flexion in the lower limb. On the basis of the motricity index, we computed a modified motricity index score, reflecting the *upper contralateral limb motor activity*, corresponding to (100 - motricity index) (Table 1). This lateralized score provides a continuous range of values from complete motor impairment (100) to normal motricity (0) of the contralesional upper limb. Scores 1-34 indicated moderate contralateral motor impairment; 35-100 severe impairment (see [29]). The score sign was set to (+) for right brain damage (left motor impairment) and to (-) for left brain damage (right motor impairment); thus -100 indicates exclusive left upper limb activity, 0 shows balanced right-left activities, and +100 indicates exclusive right upper limb activity.

Somatic and visual perception were assessed by asking patients to detect: (1) tactile stimuli on each hand and (2) visual stimuli consisting in movements of the examiner’s fingers in the visual quadrants [25,30]. Ten single and ten double symmetrical and simultaneous stimuli were presented. Scores range from 0 (no deficit) to 3 (less than 4 single stimuli reported for each limb).

### Assessment of Motor Neglect

#### a. Clinical scale and tea preparation task

We used a clinical scale [15] based on the observation of patients’ spontaneous patterns of behavior, such as: limb positioning, symmetry of the posture, presence/absence of a placing reaction, hand gesturing during speaking, arm swing during walking, underutilization, hypometria, bimanual activities, and ability to catch an object. MN was also assessed by asking patients to prepare tea [15] and video-recording their performance.

#### b. Differential actigraphy

After clinical examination, we asked patients to wear accelerometer wearable wristwatch-like accelerometers (Texas Instruments eZ-Chronos eZ430) on both wrists during 24 hours. Accelerometers were equipped with software developed in-house [17]. Patients were unaware of the aim of the study. They were instructed to continue their ordinary life, and to avoid removing the device during the recording time, with the single exception of activities involving immersion in water; these events were noted. After the recording period, we computed a score of actigraphic asymmetry, which quantifies the overall unbalance between left and right upper limbs [17]. The theoretical range of the actigraphy score ranges from -100 for exclusive left upper limb activity, to 0 for perfectly balanced right-left activities, to +100 for exclusive right upper limb activity. In order to identify patients with abnormal asymmetries in spontaneous motor behavior, we established cut-off values based on the performance of the healthy participant group tested by Rabuffetti et al. [17], by using the Crawford statistical approach [31,32]. The resulting cut-off score was 25.26 in absolute value.

### Neuroimaging study

MRI data was acquired on two different scanners with similar protocols: a 1.5 Tesla (Siemens Magnetom Avanto, Erlangen, Germany) with a 12-channel head matrix coil at IRCCS Don Carlo Gnocchi in Milan and a 3-Tesla Ingenia Philips (Philips Medical Systems, Erlangen, The Netherlands) with a 16 channels head matrix coil at the Centre GIE Imagerie Sud, Centre Hospitalier Lyon Sud. Both protocols included the following sequences: T1-weighted, T2 FLAIR and diffusion-weighted (DW) images acquired for each patient. Brain MRI scans included T13D anatomical SPGR (spoiled gradient recalled) with the following characteristics: (1) TR/TE/T1 = 7164/3124/380 ms; flip angle = 15°, matrix size = 0, 288, 256, 0; voxel resolution = 0.5 × 0.5 × 1.2 mm^3^; (2) TR/TE/TI = 1900/3.37/1.1 ms, flip angle = 15°, matrix size = 192 × 256, voxel resolution = 1 × 1 × 1 mm^3^.

Lesion masks were delineated on the original 3D images. Images were then normalized to the Montreal Neurological Institute (MNI) template using Statistical Parametric Mapping (SPM 8) (http://www.fil.ion.ucl.ac.uk/spm/software/spm) running under Matlab (Mathworks Inc., Natick, USA; www.mathworks.com/matlabcentral). The lesion extent was then segmented for each subject on normalized images by using the MRIcron software (http://www.mccauslandcenter.sc.edu/mricro/mricron/). Regions of interest (ROI) thus obtained were used in the subsequent analyses in MRIcron software for conventional lesion density plots.

### Data Availability

Data is available upon request.

## RESULTS

Table 1 shows patients’ clinical details. Table 2 presents each patient’s scores on visual and personal neglect, anosognosia and gaze orientation. All the patients obtained normal ipsilesional motricity index scores for their upper limbs. The modified motricity index score, which clinically estimates asymmetries of motricity on command, identified 19 patients with symmetric upper limbs motor ability, 4 patients with moderate motor asymmetry and 12 patients with severe motor asymmetry (Table 1).

### Motor neglect assessment

#### a. Clinical scale and tea preparation task

Ten patients showed clinical signs of motor neglect, of whom six had pathological performance on the tea task (Table 3). Three of these patients (P4, P24, P30) had signs of “pure” MN, in the absence of sensorimotor deficits, or of signs of visual and personal neglect.

**Table 3.**
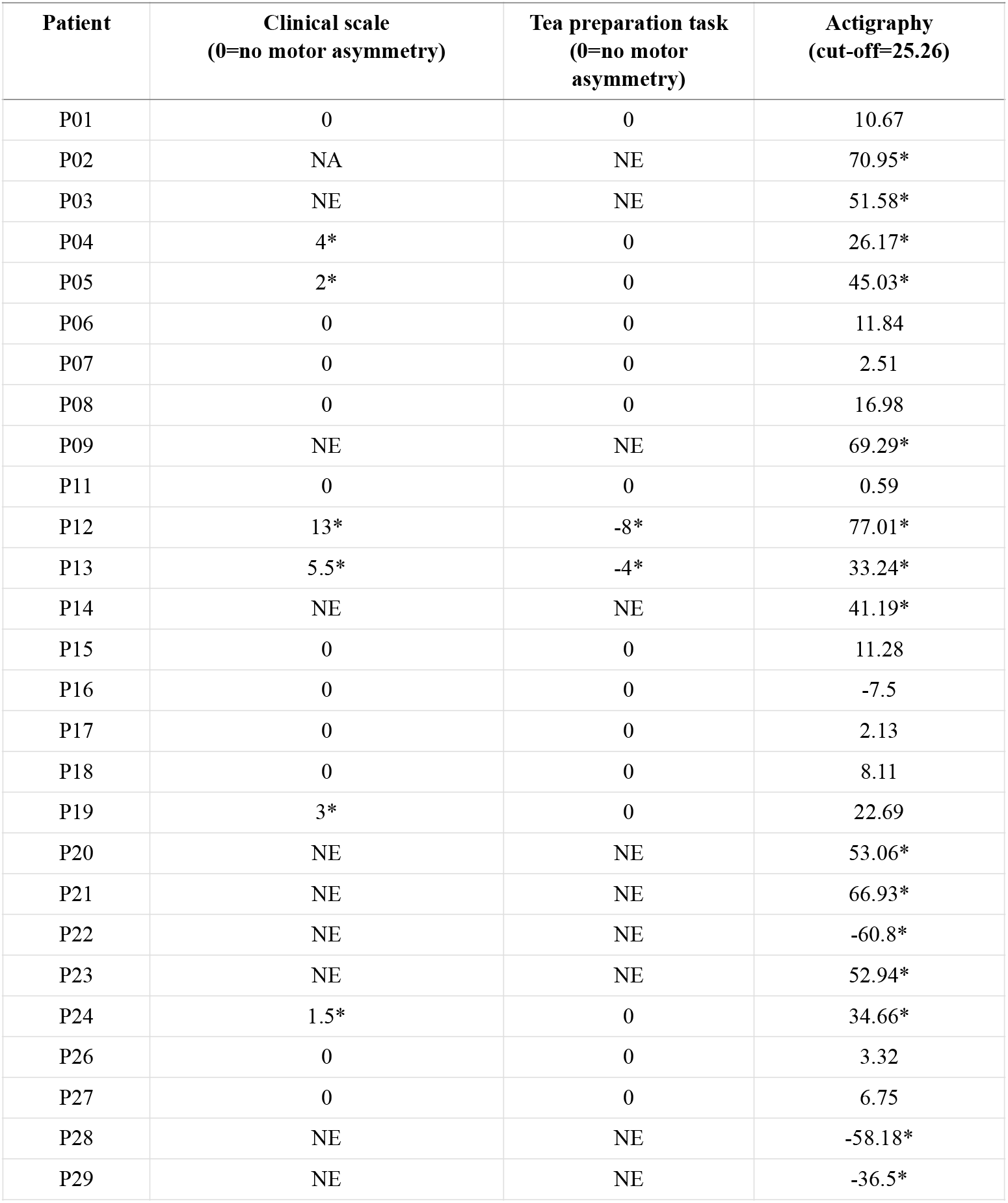
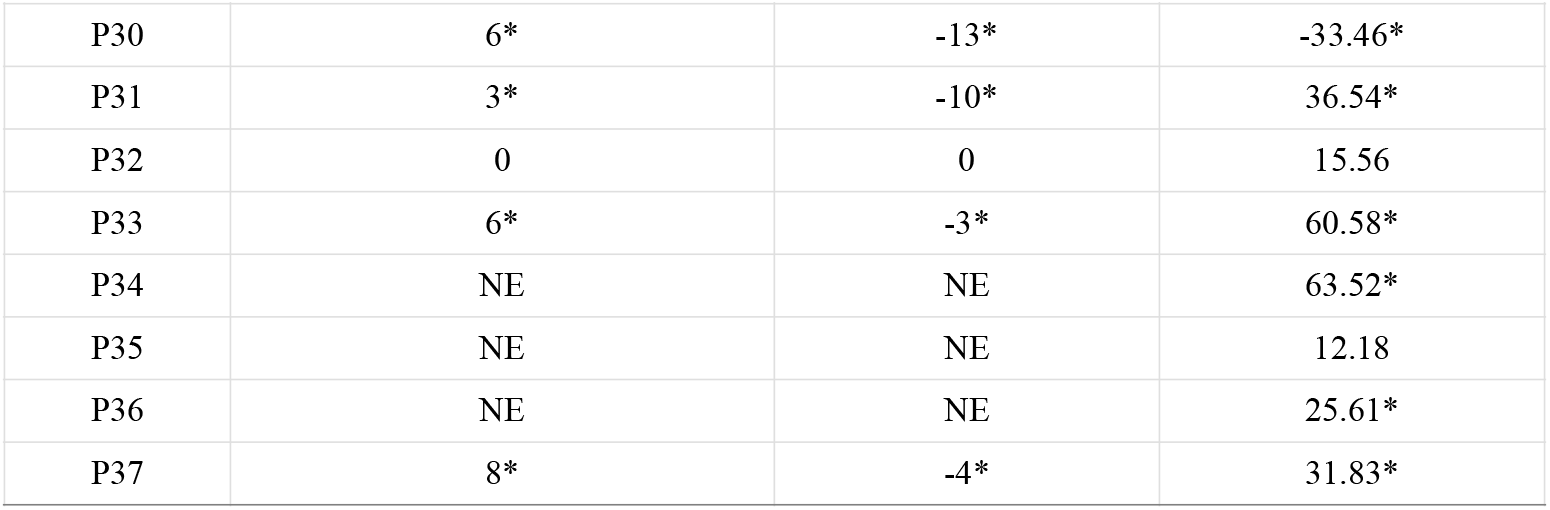
Assessment of motor asymmetry. Score obtained by each patient on the Clinical scale (range, 0 [no asymmetry in spontaneous motricity] - 20 [extremely severe MN], see [15]), the Tea preparation task (the higher the score, the more severe the motor asymmetry, see [15]), and differential actigraphy. NE, not evaluable; NA, not available. *, pathological score indicating asymmetric spontaneous motor behavior.

| **Patient** | **Clinical scale**<br>**(0=no motor asymmetry)** | **Tea preparation task**<br>**(0=no motor**<br>**asymmetry)** | **Actigraphy**<br>**(cut-off=25.26)** |
| --- | --- | --- | --- |
| P01 | 0 | 0 | 10.67 |
| P02 | NA | NE | 70.95* |
| P03 | NE | NE | 51.58* |
| P04 | 4* | 0 | 26.17* |
| P05 | 2* | 0 | 45.03* |
| P06 | 0 | 0 | 11.84 |
| P07 | 0 | 0 | 2.51 |
| P08 | 0 | 0 | 16.98 |
| P09 | NE | NE | 69.29* |
| P11 | 0 | 0 | 0.59 |
| P12 | 13* | -8* | 77.01* |
| P13 | 5.5* | -4* | 33.24* |
| P14 | NE | NE | 41.19* |
| P15 | 0 | 0 | 11.28 |
| P16 | 0 | 0 | -7.5 |
| P17 | 0 | 0 | 2.13 |
| P18 | 0 | 0 | 8.11 |
| P19 | 3* | 0 | 22.69 |
| P20 | NE | NE | 53.06* |
| P21 | NE | NE | 66.93* |
| P22 | NE | NE | -60.8* |
| P23 | NE | NE | 52.94* |
| P24 | 1.5* | 0 | 34.66* |
| P26 | 0 | 0 | 3.32 |
| P27 | 0 | 0 | 6.75 |
| P28 | NE | NE | -58.18* |
| P29 | NE | NE | -36.5* |
| P30 | 6* | -13* | -33.46* |
| P31 | 3* | -10* | 36.54* |
| P32 | 0 | 0 | 15.56 |
| P33 | 6* | -3* | 60.58* |
| P34 | NE | NE | 63.52* |
| P35 | NE | NE | 12.18 |
| P36 | NE | NE | 25.61* |
| P37 | 8* | -4* | 31.83* |

#### b. Differential actigraphy

Wearing the accelerometers was well tolerated; no patient reported any complaints concerning the procedure. Figure 1 displays representative 24-hour activity profiles of the left and the right upper limbs in a patient (P7) without substantial asymmetries, and in a patient (P33) with motor neglect.

**FIGURE 1.**
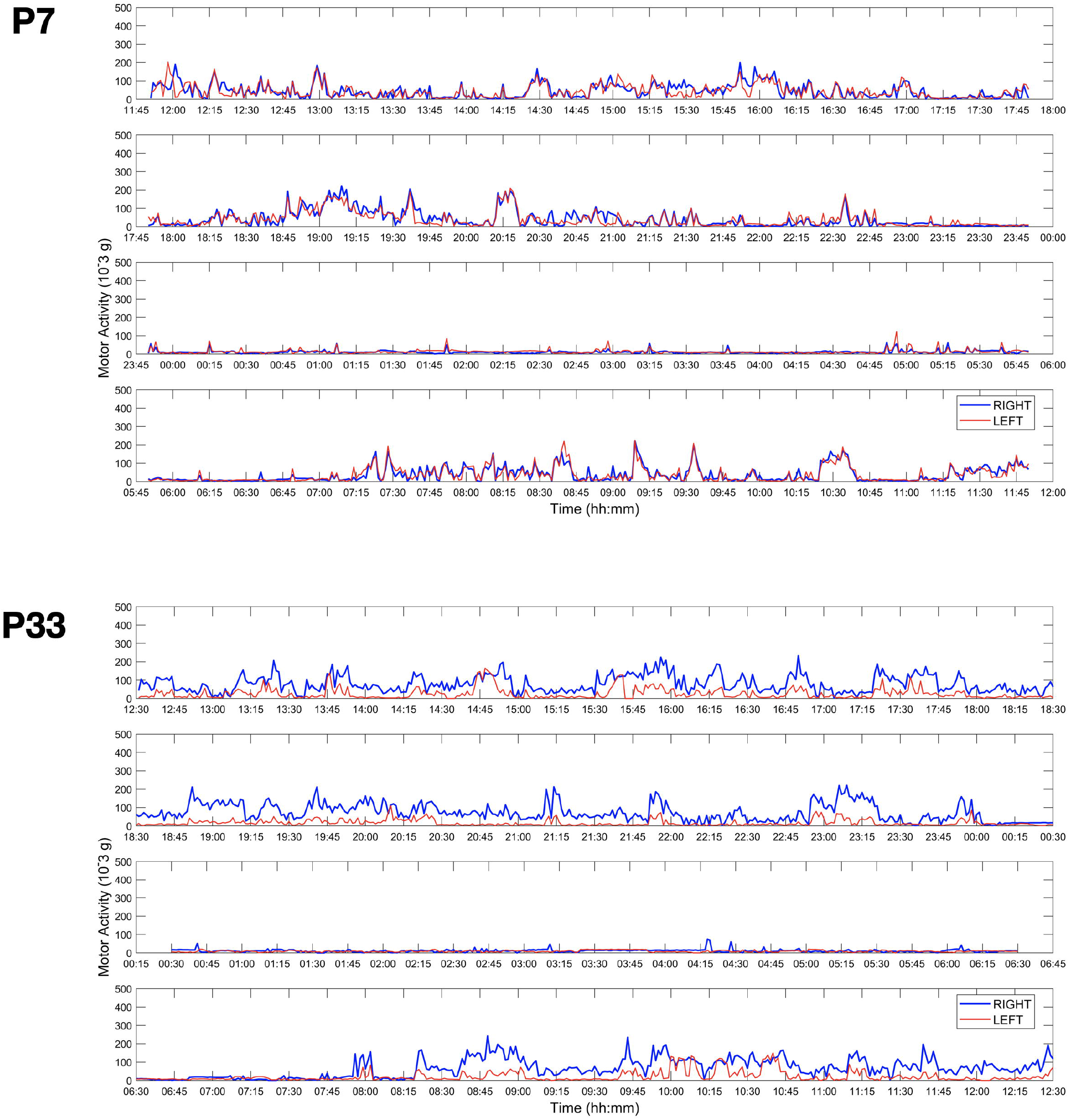
Representative examples of 24-hour activity profiles for two patients. Red, left upper limb motor activity profile; blue, right upper limb profile. Patient 7 shows normal (symmetrical) performance; Patient 33 displays a substantial right-left asymmetry, indicating motor neglect.

Fourteen patients showed symmetrical 24-hour activity profiles (see Table 3 and the central panel in Figure 2), indicating absence of MN. The remaining 21 patients showed motor asymmetries, favouring the right upper limb in 17 patients with right hemisphere damage (right-side panels in Figure 2), and the left limb in 4 patients with left brain damage (Figure 2, left-sided panels), suggesting the presence of unilateral elementary motor disorders, or of MN.

**FIGURE 2.**
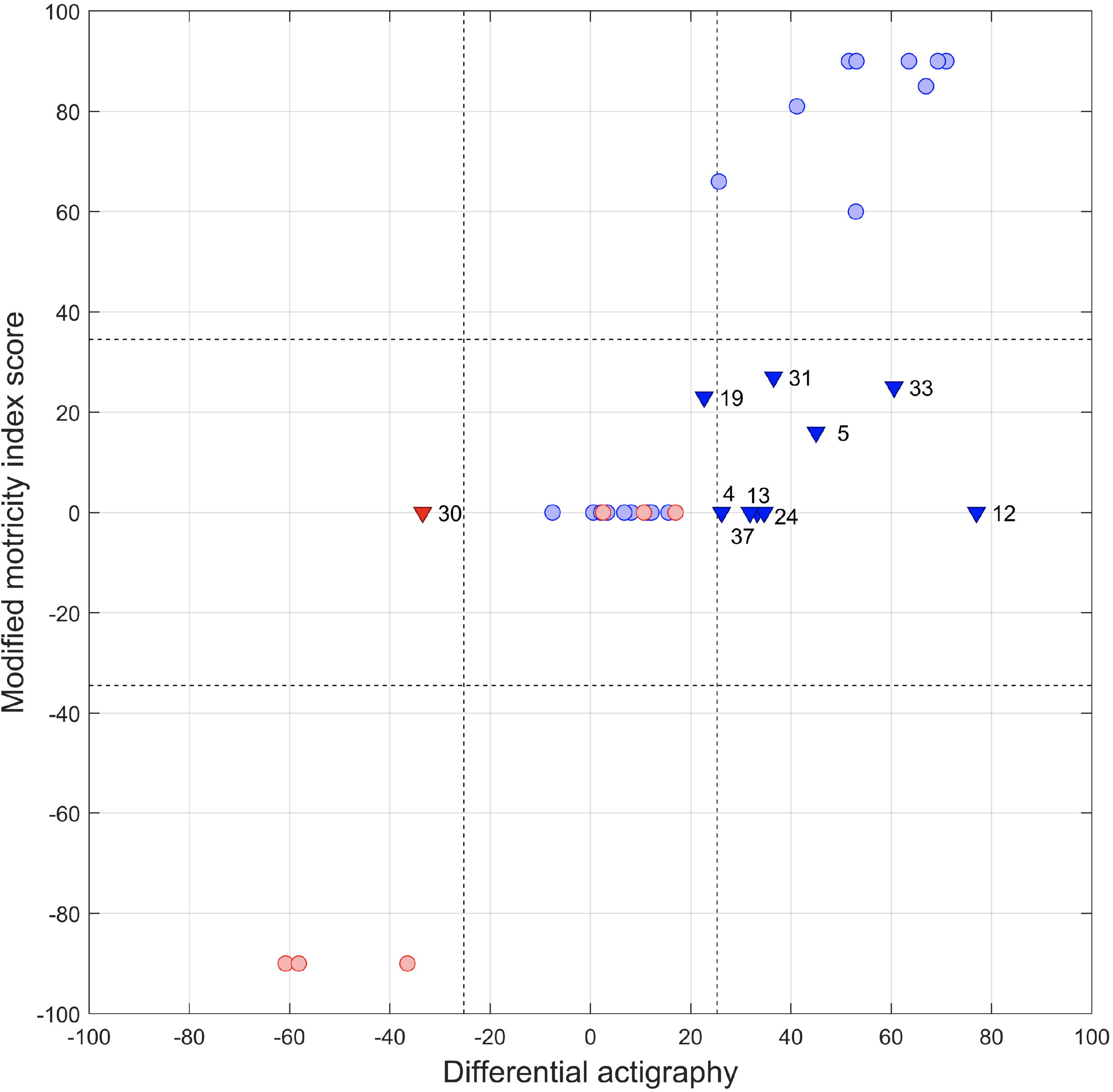
Scatterplot showing individual patients’ asymmetries in upper limb motor performance. Datapoints show each patient’s motor performance, spontaneous (differential actigraphy, x axis), and on command (modified motricity index, y axis). Positive values represent rightward asymmetry; negative values indicate leftward asymmetry. Vertical dashed lines show cutoff values for differential actigraphy; horizontal dashed lines represent conventional cutoff values for asymmetries in motor index. Red: left hemisphere damage; blue, right hemisphere damage. Triangles, patients with clinically diagnosed motor neglect (patient numbers correspond to numbers in the Tables); circles, patients without motor neglect. The upper right quadrant and the lower left quadrant include patients with contralesional hemiplegia, resulting in pathological asymmetries on both indexes.

Consideration of motricity on command (as indexed by the modified motricity index score) allowed us to distinguish between elementary motor disorders and genuine MN. Eight patients with right hemisphere damage and one patient with left hemisphere damage showed symmetrical motricity on command, but obtained asymmetrical scores on differential actigraphy (see Table 3 and the right- and left-sided central panels in Figure 2). This dissociation is typical of MN. All of these patients also had pathological scores on the clinical MN scale of Migliaccio et al. [15]. A single patient (P19) had clinical MN with borderline actigraphy score (+22.69, against a cutoff score of 25.26).

We observed the following additional patterns in our sample: (1) Relatively “pure” MN, with symmetric motricity index and asymmetric actigraphy, and no signs of visual or personal neglect (P4, P24 and P30); (2) MN associated with signs of visual and personal neglect (P12, P13), or somatosensory impairment (P37), but with symmetrical motricity index; (3) MN associated with mildly asymmetric motricity index and visual and personal neglect signs (P5, P31, P33).

### Lesion location

MRI scans were available for 25 right brain-damaged patients. Figure 3 shows lesion location in MN and non-MN patients. We labelled and quantified lesions in grey and white matter by using the Automatic Anatomical Labelling [33], and the Natbrainlab atlas [34], respectively (Table 4). In MN patients, lesional patterns were heterogenous, with most lesions encroaching upon the cortico-spinal tract and the fronto-parietal and fronto-occipital white matter bundles.

**Table 4.** Anatomical data of patients with motor neglect. Percentage of lesions was assessed by using the Automatic Anatomical Labelling [33] and Natbrainlab [34] templates. Only lesions > 10 voxels are reported here. All the patients in this table had unilateral right hemisphere lesions. #, patients with “pure” MN; CC, corpus callosum; CPC, cortico-ponto-cerebellar fibres; CS, cortico-spinal tract; IC, internal capsule; IFOF, inferior fronto-occipital fasciculus; ILF, inferior longitudinal fasciculus; OR: optic radiations.

| Patient | Grey matter lesions<br>(% lesioned voxels) | White matter lesions<br>(% lesioned voxels) |
| --- | --- | --- |
| P4# | Precentral gyrus (0.01) | CPC (0.5), CS (2.5), IC (0.2) |
| P12 | Insula (0.3), Putamen (5.8) | CPC (0.4), CS (0.6), IC (0.7) |
| P13 | Insula (0.2), Putamen (4.8), Caudate (0.2), Thalamus (0.3) ,<br>Superior (0.4) and Middle (0.6) temporal gyri | Anterior (0.6) and posterior (3.3)<br>segments of the arcuate fasciculus,<br>CPC (0.9), CS (1.8), IC (0.9), OR<br>(2.2) |
| P19 | Precentral (8.9) and Postcentral (1.68) gyri, Middle frontal<br>gyrus (0.1), Rolandic operculum (2.49), Insula (1.4),<br>Supramarginal gyrus (0.6), Heschl's gyrus (5.7), Superior<br>temporal gyrus (2.3) | Anterior segment of the arcuate<br>fasciculus (13.2), CS (0.1) |
| P24# | Insula (0.1), Putamen (10.6) | IFOF (1.7), OR (9.6) |
| P31 | Insula (0.2), Amygdala (1.7), Caudate (0.1), Putamen (16.3),<br>Pallidum (4.1) | Anterior segment of the arcuate<br>fasciculus (1.1), CPC (0.8), CS (5.5),<br>IFOF (2), IC (1.2) |
| P33 | Precentral (4.5) and Postcentral (0.1) gyri, Superior (33.8),<br>Middle (57.8) and Inferior (36.4) frontal gyrus , Superior<br>(44.6), Middle (36.1) and Inferior (4.3) orbital frontal<br>regions, Olfactory cortex (6.3), Medial frontal superior<br>(44.4), medial orbital frontal (78), Gyrus rectus (30.4), Insula<br>(4.9), Cingulate anterior (23.3), Amygdala (2.9), Caudate<br>(17.9), Putamen (63.8), Pallidum (11.7), Superior (22.8),<br>Middle (3.3) and Inferior (0.1) temporal gyri, Superior (9.7)<br>and Middle (1.2) temporal pole | Anterior commissure (1.3), Anterior<br>segment of the arcuate fasciculus<br>(1.3), Cingulum (8.2), CC (11.4),<br>CPC (1.1), CS (8.4), ILF (0.1), IFOF<br>(14.8), IC (15.1), OR (1.5) |
| P37 | Cuneus (3.6), Precuneus (0.6), Lingual (5.6) and Fusiform<br>(12.9) gyri, Superior (6), Middle (12.3) and Inferior (67.6)<br>occipital gyri, Superior parietal lobule(0.8), Middle (1.4) and<br>Inferior (20.1) temporal gyri, Cerebellum (42) | CC (0.2), ILF (2.2), IFOF (5.3) |

**FIGURE 3.**
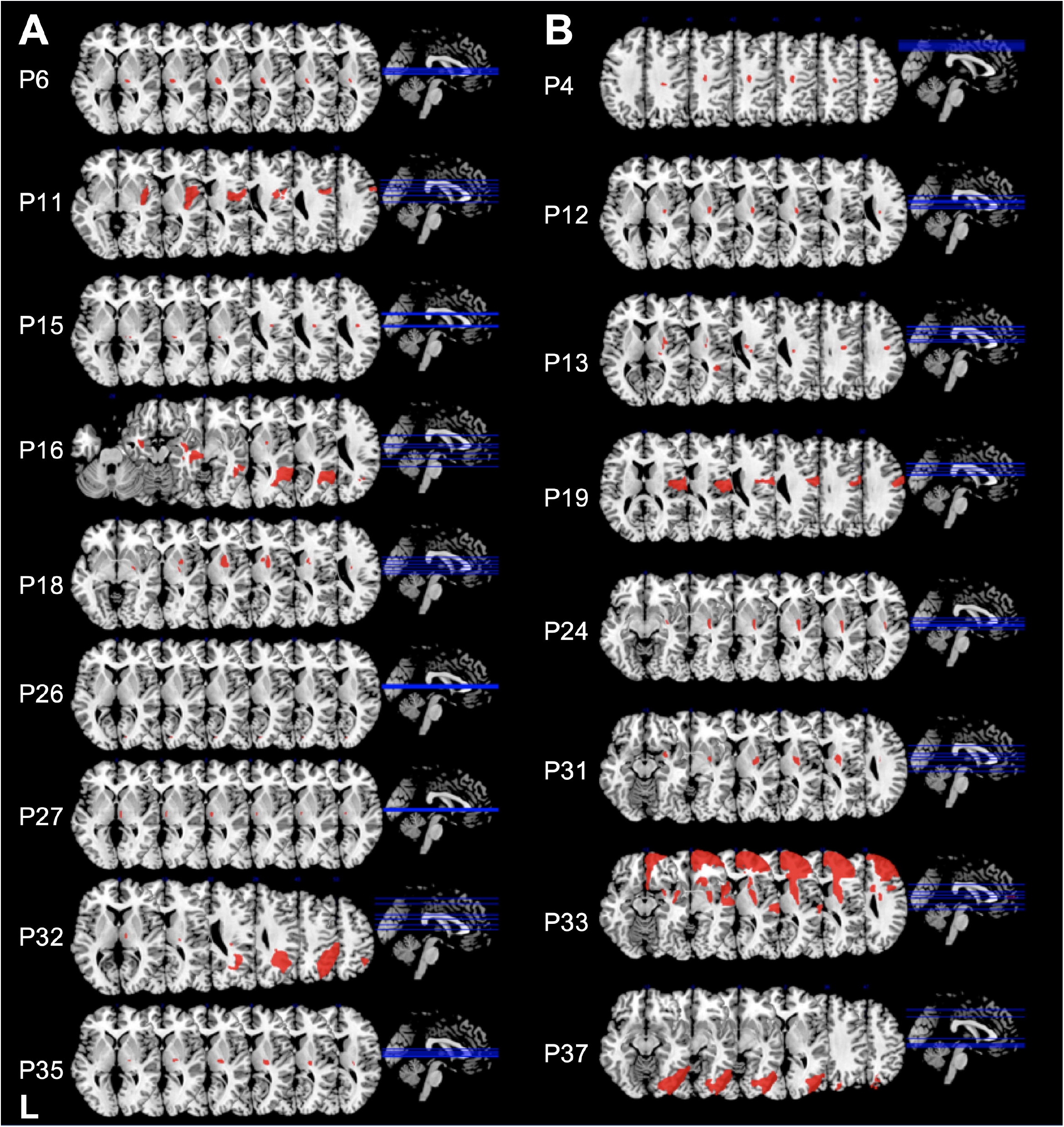
Lesion location in right brain-damaged patients. (A) Patients with no lateralized motor deficits. (B) Patients showing signs of motor neglect. Lesion location in patients without lateralized motor deficits included thalamus, putamen, insula, parieto-temporal regions and internal capsule. Lesion location in MN patients predominantly included putamen, insula and internal capsule.

## DISCUSSION

Here we present 24-hour differential actigraphy as a tool to objectively assess and quantify motor neglect in brain-damaged patients. Our technique resolves a major issue of diagnosis of MN, which is only evident for spontaneous movements, whereas movements on command are normally executed. At present, clinical diagnosis of MN is only observational. It depends on subjective clinical assessment, which requires a substantial amount of training. By contrast, administration and automated analysis of differential actigraphy can easily be accomplished with minimal training. Our results were remarkably consistent with the outcome of qualitative clinical observation, with the single exception of P19, who had clinical signs of MN and obtained a borderline asymmetry score on actigraphy. A further, important advantage of actigraphy over clinical observation is the ability to provide detailed quantitative measures of asymmetries of spontaneous motor behavior. Differential actigraphy can thus be used not only for initial diagnosis, but also for patient follow-up, to evaluate the evolution of MN and the effects of rehabilitation.

Differential actigraphy proved to be more sensitive than the tea preparation task used in previous research [15]. Only six patients of the present sample showed signs of MN on the tea preparation task, perhaps because knowledge of being videotaped made their motor behavior less spontaneous and more controlled. Actigraphy showed asymmetric motor behavior in all these patients, plus three more.

The combined assessment of visual and personal neglect in our sample enabled us to evaluate the possibility of dissociated patterns of performance. Our results confirmed that MN can occur in the absence of signs of visual neglect [2], or of personal neglect [35]. The dissociation from personal neglect is of theoretical relevance, because it challenges the hypothesis that all MN patients simply do not pay attention to their contralesional limbs.

In a subset of 25 right-brain damaged patients, we explored the lesional correlates of MN. Lesion location was heterogeneous, but frequently involved the white matter, including cortico-spinal tracts and long-range fronto-parietal and fronto-occipital fascicles. The implication of medial fronto-parietal networks is consistent with their role in the initialization of a voluntary action (whether, how and when to act, see [36]). However, only three patients in our sample had lesions in or near the supplementary motor area, and only one patient had a cingulum disconnection, in contrast with previous studies [15,16]. Damage to fronto-occipital connections, which convey top-down influence from prefrontal cortex on posterior visual areas, and whose damage has been associated with visual neglect [37–40], is more difficult to relate to MN. Given their length, fronto-occipital connections are relatively likely to be affected by brain damage, and might thus represent an “innocent bystander” in the case of MN. Lesion patterns in patients presenting “pure” MN (P4, P24, P30) implicated the cortico-spinal tracts and the putamen. The substantial variability of lesion location may reflect functional heterogeneity of motor neglect. However, given the limited patient sample, the specificity of these anatomical findings awaits confirmation. Another limitation of our anatomical analysis is that patient assessment and neuroimaging were performed at variable time intervals among patients. Therefore, it is difficult to compare behavioral patterns and lesions between patients at acute/subacute phase and patients in the chronic phase.

In conclusion, our findings indicate that differential actigraphy, together with appropriate analysis methods, offers a convenient, cost-effective, and relatively automatized procedure to follow-up motor behavior in neurological patients, and to assess the effects of rehabilitation. Furthermore, actigraphy provides neuroscientists with a suitable tool to study the neural bases of spontaneous movements in neurological patients. Differential actigraphy should be included in the routine evaluation and follow-up of motor abilities in stroke survivors.

## Data Availability

Data is available from Dr Chiara Pagliari upon request.

## ACKNOWLEDGMENTS

The work of PB is supported by Agence Nationale de la Recherche through ANR-16-CE37-0005 and ANR-10-IAIHU-06. CP was partially funded by the Italian Ministry of Health, Ricerca Corrente 2018-2020.

## APPENDIX 1

### AUTHORS

APPENDIX 1: AUTHORS
| Author | Location | Contribution |
| --- | --- | --- |
| Monica N. Toba PhD | University of Picardy<br>Jules Verne, Amiens,<br>France | Performed the analyses; interpreted the data; drafted the manuscript for intellectual content |
| Chiara Pagliari PhD | Fondazione Don Carlo Gnocchi, Milano, Italy | Acquired the data, performed the analyses; drafted the manuscript for intellectual content |
| Marco Rabuffetti<br>DrEng | Fondazione Don Carlo Gnocchi, Milano, Italy | Conceived the methodological approach; performed the statistical analyses; drafted the manuscript for intellectual content |
| Norbert Nighoghossian<br>MD | Hospices Civils de Lyon, France | Contributed to data acquisition; revised the manuscript for intellectual content |
| Gilles Rode MD PhD | Hospices Civils de Lyon, France | Contributed to data acquisition; revised the manuscript for intellectual content |
| François Cotton MD | Hospices Civils de Lyon, France | Contributed to data acquisition; revised the manuscript for intellectual content |
| Lucia Spinazzola PhD | Humanitas Mater Domini Hospital, Castellanza, Italy | Contributed to data acquisition; revised the manuscript for intellectual content |
| Francesca Baglio MD | Fondazione Don Carlo Gnocchi, Milano, Italy | Contributed to data acquisition; revised the manuscript for intellectual content |
| Raffaella Migliaccio<br>MD PhD | Paris Brain Institute, ICM, France | Reviewed analyses; revised the manuscript for intellectual content |
| Paolo Bartolomeo MD<br>PhD | Paris Brain Institute, ICM, France | Conceived and coordinated the study; drafted the manuscript for intellectual content |

